# Development and validation of personalised risk prediction models for early detection and diagnosis of primary liver cancer among the English primary care population using the QResearch^®^ database: research protocol and statistical analysis plan

**DOI:** 10.1101/2022.01.24.22269755

**Authors:** Weiqi Liao, Peter Jepsen, Carol Coupland, Hamish Innes, Philippa C. Matthews, Cori Campbell, Eleanor Barnes, Julia Hippisley-Cox, DeLIVER consortium

**Affiliations:** Nuffield Department of Primary Care Health Sciences, University of Oxford, Oxford, UK; Department of Hepatology and Gastroenterology, Aarhus University Hospital, Aarhus, Denmark; Division of Primary Care, School of Medicine, University of Nottingham, Nottingham, UK; School of Health and Life Sciences, Glasgow Caledonian University, Glasgow, UK; The Francis Crick Institute, London, UK; University College London, London, UK; Nuffield Department of Medicine, University of Oxford, Oxford, UK

**Keywords:** liver cancer, hepatocellular carcinoma (HCC), cholangiocarcinoma, risk prediction model, early detection, diagnosis, symptom, comorbidity

## Abstract

**Background and research aim:** The incidence and mortality of liver cancer have been increasing in recent years in the UK. However, liver cancer is still under-studied. The Early <u>De</u>tection of Hepatocellular <u>Liver</u> Cancer (DeLIVER-QResearch) project aims to address the research gap and generate new knowledge to improve early detection and diagnosis of **primary** liver cancer from general practice and at the population level. There are three research objectives: (1) to understand the current epidemiology of primary liver cancer in England, (2) to identify and quantify the symptoms and comorbidities associated with liver cancer, and (3) to develop and validate prediction models for early detection of liver cancer suitable for implementation in clinical settings.

**Methods:** This population-based study uses the QResearch® database (version 46) and includes patients aged 25-84 years old and without a diagnosis of liver cancer at the cohort entry (study period: 1 January 2008 to 31 December 2020). The team conducted a literature review (with additional clinical input) to inform the inclusion of variables for data extraction from the QResearch database. A wide range of statistical techniques will be used for the three research objectives, including descriptive statistics, multiple imputation for missing data, conditional logistic regression to investigate the association between the clinical features (symptoms and comorbidities) and the outcome, fractional polynomial terms to explore the non-linear relationship between continuous variables and the outcome, and Cox regression for the prediction model. We have a specific focus on the 1-year, 5-year, and 10-year absolute risks of developing liver cancer, as risks at different time points have different clinical implications. The internal-external validation approach will be used, and the discrimination and calibration of the prediction model will be evaluated.

**Discussion:** The DeLIVER-QResearch project uses large-scale representative population-based data to address the most relevant research questions for early detection and diagnosis of primary liver cancer in England. This project has great potential to inform the national cancer strategic plan and yield substantial public and societal benefits.

**Medical Subject Headings (MeSH):** Liver Neoplasms; Carcinoma, Hepatocellular; Cholangiocarcinoma; Clinical Decision Rules; Early Detection of Cancer; Early Diagnosis; Symptom Assessment; Comorbidity

## Introduction and research background

According to the most recent statistics from Cancer Research UK, liver cancer represents the 18^th^ most common cancer in the UK, accounting for 2% of all new cancer cases in 2017. However, it is the 8^th^ most common cause of cancer death, accounting for 3% of all cancer deaths in 2018 [1]. Deaths due to liver cancer have increased by around 50% in the last decade [2]. The age-standardised incidence and incidence-based mortality rates of primary liver cancer increased during 1997-2017, particularly notable in hepatocellular carcinoma (HCC) [3]. The rapid increase in incidence and mortality rates is a public health concern, and a burden to health and social care. Compared with other cancers, the prognosis of liver cancer is poor, with respective 1- and 5-year survival estimates of only 38.1% and 12.7% in England during 2013-2017 [1]. Early diagnosis is associated with better survival. When it is diagnosed at its earliest stage, 78% of people can survive for one year or longer, compared with 20% when diagnosed at the latest stage [1, 4, 5].

A better understanding of the clinical features that indicate the development and progression of liver cancer would improve prompt referrals for investigation and intervention. Risk prediction models to estimate an individual patient’s future risk of developing liver cancer would enhance early detection and diagnosis (ED&D) of liver cancer in the English population, and could inform screening and prevention strategies. For example, individuals at high risk can be identified for active surveillance using liver ultrasonography and serum alpha-fetoprotein (AFP) test [6], which could help to shift the diagnosis of liver cancer towards earlier stages, when more treatment options are available and patients have a greater chance of better outcomes. This would contribute to the national cancer strategic plan for better cancer survival outcomes [7]. All of these are the motivations to conduct the DeLIVER-QResearch project, which is part of an initiative funded by Cancer Research UK to early <u>de</u>tect hepatocellular <u>liver</u> cancer (DeLIVER), project website: www.deliver.cancer.ox.ac.uk.

The QResearch team has conducted previous work on developing the QCancer prediction models to estimate future risk of lung, colorectal, gastro-oesophageal, pancreas, blood and renal tract cancers for both men and women, breast, cervical, and ovarian cancers for women, and prostate and testicular cancers for men [8]. The QCancer algorithms have been implemented in the NHS computer system as decision support tools for GPs. An online risk calculator is also available at https://www.qcancer.org/, free for public use. However, liver cancer is still not part of the QCancer model family yet. Therefore, this project will address this gap.

### Research aim and objectives

This study aims to generate new knowledge for early detection and diagnosis of primary liver cancer from the English population, with a special interest in HCC. There are three research objectives in the DeLIVER-QResearch project:

1. To understand the current epidemiology of people diagnosed with liver cancer in England, including the incidence, route to diagnosis, cancer stage and histology, treatments, survival duration, and the causes of death;
2. To explore and characterise the symptom and comorbidity profile for patients diagnosed with liver cancer, compared with those without liver cancer;
3. To develop and validate personalised prediction models for estimating the risk of patients getting a diagnosis of liver cancer in the next 5 or 10 years using primary care electronic health records (EHRs), which could identify patients at the highest risk who will benefit the most from active surveillance and early clinical intervention.

## Methods

### Study designs

An open cohort study will be used for research objectives 1 and 3, and a nested case-control study (within the same cohort) for research objective 2.

### Data source – the QResearch® database

Routinely collected EHRs linked to the QResearch database (version 46) will be the main data source for this study. QResearch is a large consolidated database with anonymised EHRs of over 35 million patients from 1800+ general practices using the Egton Medical Information Systems (EMIS) spread across England. The database includes patients who are currently registered with the practices as well as historical patients who may have left or died. Historical records date back to 1989 with linked data on all practices since 1998. Patients’ primary care records are linked with other national datasets, such as the Hospital Episode Statistics (HES, secondary care data, including inpatient, outpatient, accident and emergency (A&E), and critical care), death registration data (up to 15 causes of death) from the Office for National Statistics (ONS), and cancer registration data from Public Health England (PHE).

The team conducted a rapid literature review (including the NICE guidelines) and had clinical input from specialists (hepatology, primary care, infectious diseases) to inform the inclusion of variables and prepare code lists to extract data from the QResearch database. Two patient representatives reviewed the lists of symptoms and comorbidities and shared their experiences and disease trajectories with the lead researcher. We prepared Read/SNOMED-CT code lists to extract events from the GP records, ICD-10 code lists for diagnosed diseases in the HES, cancer registry and death records, and OPCS code lists for interventions and procedures conducted in NHS hospitals. Tables 1 and 2 summarise the variables we requested for the three research objectives. Some variables may not be significantly associated with the outcome after the analysis. However, we do not want to miss any potential association. Therefore, the variables are included as broadly and comprehensively as possible in the project set-up phase.

**Table 1.** General variables for this study

| **Data source** | **Categories** | **Variables** |
| --- | --- | --- |
| **GP record** | Demographics | Age, sex, ethnicity, socioeconomic deprivation (Townsend quintile), geographical regions in England |
|  | Lifestyle | Body mass index (BMI, continuous variable), smoking and drinking status and intensity (with units) – longitudinal data available for all variables |
| **HES** |  | Diagnoses and treatments in the outpatient appointments and hospital admissions |
| **Cancer registry (PHE)** |  | Date of liver cancer diagnosis, route to diagnosis, stage at diagnosis, cancer grade, and histology |
| **HES** | Treatments | Surgical resection of liver, liver transplantation, ablation of liver, transarterial chemoembolization (TACE), transarterial radioembolization (TARE) – OPCS codes |
| **ONS death** |  | Date of death, all causes of death (up to 15) |

Note: HES – Hospital episode statistics, ONS – Office for National Statistics, PHE – Public Health England

**Table 2.**
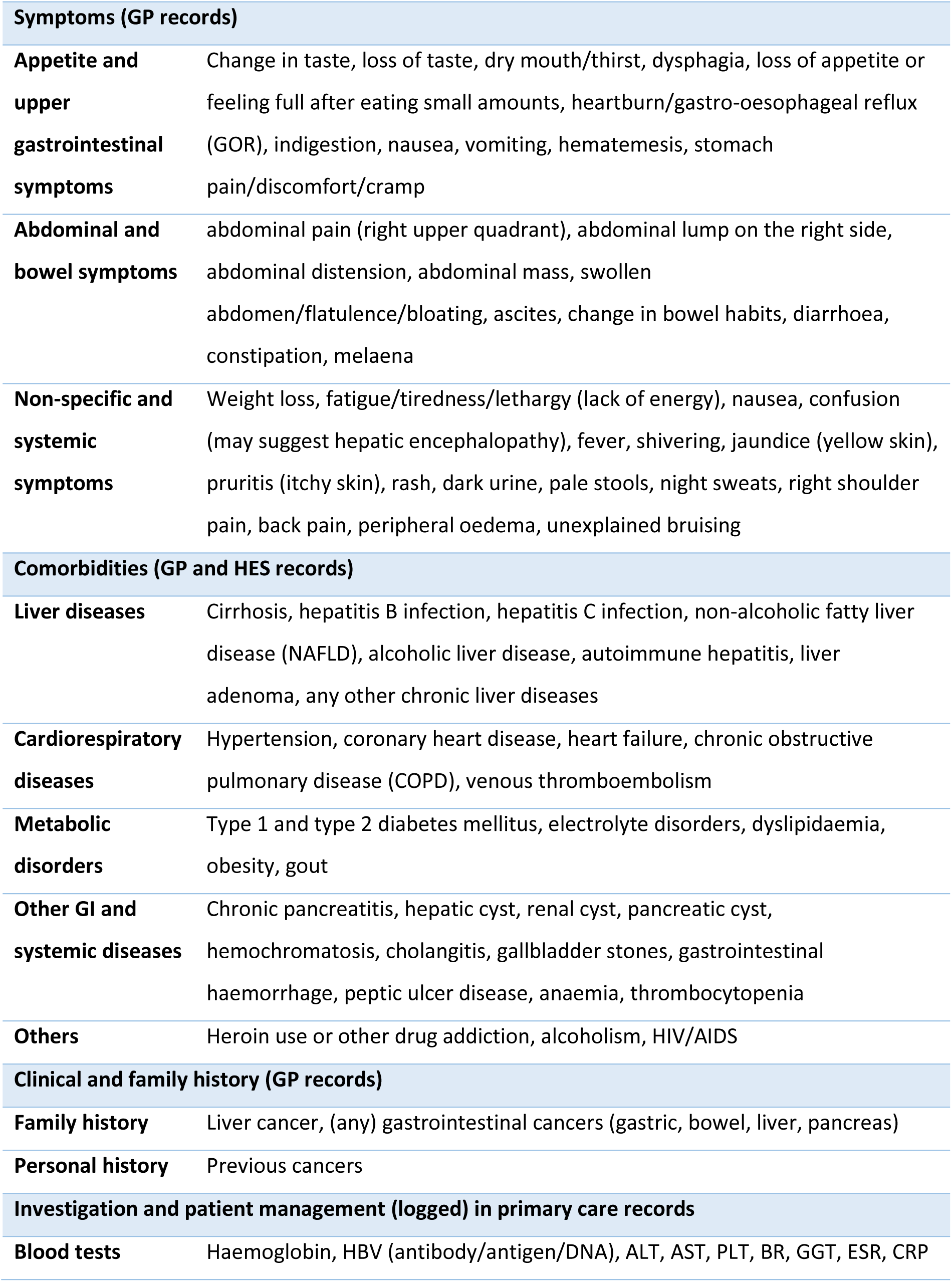
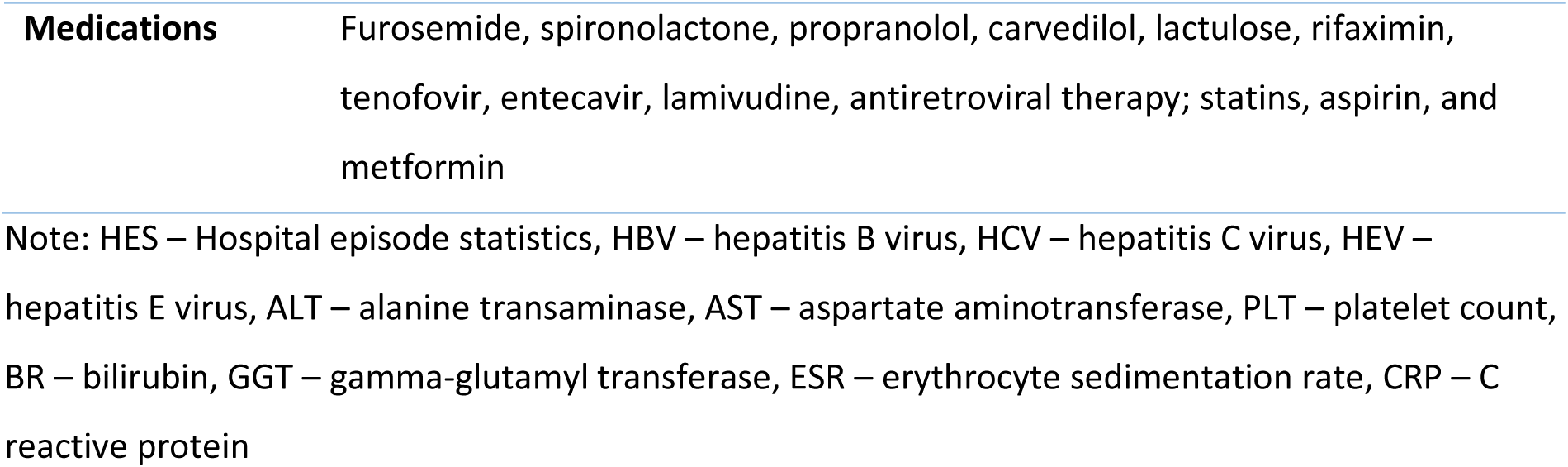
Symptoms, comorbidities and clinical characteristics relevant to liver cancer

| **Symptoms (GP records)** |  |
| --- | --- |
| **Appetite and upper gastrointestinal symptoms** | Change in taste, loss of taste, dry mouth/thirst, dysphagia, loss of appetite or feeling full after eating small amounts, heartburn/gastro-oesophageal reflux (GOR), indigestion, nausea, vomiting, hematemesis, stomach pain/discomfort/cramp |
| **Abdominal and bowel symptoms** | abdominal pain (right upper quadrant), abdominal lump on the right side, abdominal distension, abdominal mass, swollen abdomen/flatulence/bloating, ascites, change in bowel habits, diarrhoea, constipation, melaena |
| **Non-specific and systemic symptoms** | Weight loss, fatigue/tiredness/lethargy (lack of energy), nausea, confusion (may suggest hepatic encephalopathy), fever, shivering, jaundice (yellow skin), pruritis (itchy skin), rash, dark urine, pale stools, night sweats, right shoulder pain, back pain, peripheral oedema, unexplained bruising |
| **Comorbidities (GP and HES records)** |  |
| **Liver diseases** | Cirrhosis, hepatitis B infection, hepatitis C infection, non-alcoholic fatty liver disease (NAFLD), alcoholic liver disease, autoimmune hepatitis, liver adenoma, any other chronic liver diseases |
| **Cardiorespiratory diseases** | Hypertension, coronary heart disease, heart failure, chronic obstructive pulmonary disease (COPD), venous thromboembolism |
| **Metabolic disorders** | Type 1 and type 2 diabetes mellitus, electrolyte disorders, dyslipidaemia, obesity, gout |
| **Other GI and systemic diseases** | Chronic pancreatitis, hepatic cyst, renal cyst, pancreatic cyst, hemochromatosis, cholangitis, gallbladder stones, gastrointestinal haemorrhage, peptic ulcer disease, anaemia, thrombocytopenia |
| **Others** | Heroin use or other drug addiction, alcoholism, HIV/AIDS |
| **Clinical and family history (GP records)** |  |
| **Family history** | Liver cancer, (any) gastrointestinal cancers (gastric, bowel, liver, pancreas) |
| **Personal history** | Previous cancers |
| **Investigation and patient management (logged) in primary care records** |  |
| **Blood tests** | Haemoglobin, HBV (antibody/antigen/DNA), ALT, AST, PLT, BR, GGT, ESR, CRP |
| **Medications** | Furosemide, spironolactone, propranolol, carvedilol, lactulose, rifaximin, tenofovir, entecavir, lamivudine, antiretroviral therapy; statins, aspirin, and metformin |

Note: HES – Hospital episode statistics, HBV – hepatitis B virus, HCV – hepatitis C virus, HEV – hepatitis E virus, ALT – alanine transaminase, AST – aspartate aminotransferase, PLT – platelet count, BR – bilirubin, GGT – gamma-glutamyl transferase, ESR – erythrocyte sedimentation rate, CRP – C reactive protein

### Study setting and population

This is a population-based study. Patients in the QResearch database from 1 January 2008 to 31 December 2020 are eligible to be included as the study population. We use similar inclusion and exclusion criteria as those in previous studies [8-10] to develop and validate the other QCancer models. The study population will be adult patients aged between 25 and 84 years old and without a diagnosis of liver cancer at the cohort entry (1 January 2008). The age range covers the majority of patients, who are more likely to benefit from active surveillance and screening for early diagnosis of liver cancer. The included patients need to have been registered in the general practices for at least 12 months, and these practices need to have contributed to the QResearch database for a minimum of 12 months before the cohort entry date. This is to ensure complete data before cohort entry. Figure 1 shows the timeline of the dynamic patient cohort for the DeLIVER-QResearch project.

**Figure 1.**
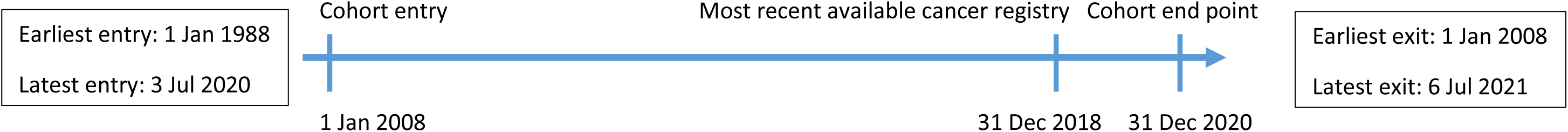
Timeline of the dynamic patient cohort for the DeLIVER-QResearch (version 46) project

### Identification of liver cancer cases

Incident primary liver cancer cases during 2008-2018 (the most recent available data from the linked cancer registry) will be identified and followed up to mid-2021 (the data flow to the most recent QResearch version). Data on treatments and outcomes (e.g. death, left cohort, still alive) for the liver cancer cases are available in the follow-up period in different datasets. Patients with secondary liver cancer (metastasis, indicated by ICD-10 codes) will be excluded, as they are not suitable for the research purposes of this project.

### Study outcomes

The primary outcome is the incident diagnosis of liver cancer recorded on any of the four aforementioned linked data sources. The date of diagnosis will be the earliest date recorded on any of the four data sources, which will be the index date in the nested case-control study. The secondary outcomes are the liver cancer subtypes (e.g. HCC, cholangiocarcinoma) and the stage at diagnosis (likely to convert into a binary variable, i.e. early vs late stage).

### Control groups in the nested case-control study

Around 80%-90% of patients have cirrhosis before being diagnosed with HCC [11]. Cirrhosis could be seen as one possible precursor and on the disease trajectory of developing HCC. Therefore, to better understand the clinical features and early signs of liver cancer, we will select two control groups (the general primary care population and patients diagnosed with cirrhosis) in the nested case-control study. The controls will be matched by age, sex, practice, and calendar year (where possible) with each cancer case using incidence density sampling [12]. Each control will be allocated an index date which is the date of cancer diagnosis in their matched case.

### Ethical approval

This study has been approved by the QResearch Scientific Committee on 8 July 2021. QResearch is a Research Ethics Approved Research Database, confirmed from the East Midlands – Derby Research Ethics Committee (research ethics reference: 18/EM/0400, project reference: OX30 DeLIVER). A dedicated webpage for this project has been created on the QResearch website https://www.qresearch.org/research/approved-research-programs-and-projects/development-and-validation-of-personalised-risk-prediction-models-for-early-detection-and-diagnosis-of-hepatocellular-carcinoma-hcc-among-the-english-population-from-primary-care/. A lay summary of this project for the public audience is available on the webpage.

## Statistical analysis plan

### General statistical methods used in multiple research objectives

#### Descriptive statistics

Descriptive statistics will be used to describe the sociodemographic (e.g. age, sex, ethnicity, Townsend quintile) and clinical characteristics (e.g. symptoms, comorbidities, cancer stage, grade, histology) of the study population, using means and standard deviations, medians and interquartile ranges (IQR), and proportions, for different types of data where appropriate. For the nested case-control study, characteristics will be described separately for cases and the two sets of controls. We will describe the temporal changes in incidence, routes to diagnosis, stage at diagnosis, and treatments for liver cancer by year during the study period.

#### Handling missing data

For symptoms, comorbidities, and medication use, the absence of information in the EHRs will be assumed that the patient did not have the health conditions or was not prescribed the medications. There may be missing data in some variables, as they were not collected and recorded in the EHRs. We will use multiple imputation with chained equations (MICE) to replace missing values for ethnicity, Townsend quintile, BMI, smoking status, and alcohol intake, with the assumption of data missing at random (MAR) [13-16]. Five imputations will be conducted, as this has relatively high efficiency [17] and is a pragmatic approach accounting for the size of the data and the capacity of the available computing power in software and the server. Rubin’s rules will be used to combine the model parameter estimates across the imputed datasets [18].

#### Variable selection and considerations for regression models

The backward elimination approach in multivariable modelling is preferred in the TRIPOD guideline [17]. Thus, we will fit the models by including all the variables initially, and then retain those having odds ratios (OR, in conditional logistic regression for research objective 2) or hazard ratios (HR, in Cox regression for research objective 3) <0.91 or >1.10 (clinical significance) for binary/categorical variables and at the statistical significance level of 0.01 (two-tailed). For previous diagnoses of other cancers, we will retain the variables at the significance level of 0.05, since some cancers are rare and may not have enough numbers. To simplify the models, we will focus on the most common health conditions and medications, and combine similar variables with comparable ORs or HRs where possible. If some variables do not have enough events to obtain point estimates and standard errors, we will combine some of these if clinically similar in nature.

### Epidemiology of primary liver cancer and cirrhosis (research objective 1)

The overall incidence rate of liver cancer and by age groups, sex, ethnicity, socioeconomic deprivation (Townsend quintiles), the ten geographical regions in England, and liver cancer subtypes (HCC, cholangiocarcinoma, other specified or unspecified liver cancer) per 100,000 person-years will be calculated. Poisson regression will be used to investigate how patient characteristics are associated with the trend over time and the variation in the incidence of liver cancer and HCC throughout the study period, where interactions between age, sex, ethnicity, and Townsend quintile will be explored. In addition, we will explore factors associated with emergency presentation, late stage at diagnosis, patients received curative treatments or any treatment, and survival duration, with a specific interest in the effects of sex, socioeconomic deprivation, and ethnicity. Considering the importance of cirrhosis on the disease trajectory of liver cancer, we will use similar methods to establish the epidemiology (e.g. incidence, prevalence, the trend over the years, and distribution in the population) of cirrhosis using EHRs before conducting the nested case-control study.

### Explore the clinical features and red-flag symptoms associated with liver cancer (research objective 2)

#### The most common symptoms and the combinations

Patients may present to their GP with a series of different symptoms. Besides each individual symptom, the frequency of the most common symptom combinations will be summarised, which would provide additional helpful information for GPs to manage patients based on symptomatic presentation and inform the development of diagnostic models in future studies.

#### The analyses for the nested case-control study

We will first explore the symptoms and comorbidities in patients’ EHRs in different timeframes, e.g. up to 3 months, 4-6 months, 7-12 months, 1-2 years, 2-3 years, and 3-5 years, before the date of diagnosis in cases, or the equivalent date in the matched controls. Multivariable conditional logistic regression will be used for the matched case-control design, adjusting for confounders (e.g. patient’s sociodemographic characteristics and lifestyle factors) and considering the interaction terms. This will determine which ‘red flag’ symptoms remain significant when other factors are taken into account in the model. After handling the missing data (subsection above), the results from the imputed datasets will be the primary estimates, but these will be compared with the estimates from two sets of sensitivity analyses, which are complete case analysis, and analyses restricted to cases and controls that have more than three years of EHRs before the index date. The analyses will be carried out separately for the two control groups (the general primary care population and patients diagnosed with cirrhosis), and the findings from the two control groups will be compared. Should the cancer staging be sufficiently complete, we will conduct further analysis to identify clinical features that can help diagnose liver cancer earlier. The reporting of the case-control study will follow the recommendations of the STROBE statement [19].

### Methodology for prediction models to estimate individual risk of developing liver cancer (research objective 3)

#### Sample size considerations

We used the criteria by Riley et al. [20] and the ‘pmsampsize’ package in R to calculate the minimum required sample size for developing a clinical prediction model. The parameters for sample size estimation for time-to-event outcome were set or assumed as follows. The previous QCancer prognostic models have around 30 predictors, we assume 50 predictors in the new models to allow more flexibility. The median duration from cohort entry to the incident diagnosis of liver cancer is about 10 years (QResearch has linked data on all practices since 1998, see the data source section above), which is suitable for the predictive period (up to 10 years). The age-standardised incidence rate (event rate) of liver cancer in the UK during 2016-2018 was 14.5 (95% CI: 14.3-14.8) per 100,000 population for men and 6.2 (95% CI: 6.0-6.3) for women (statistics from Cancer Research UK [21]). A conservative 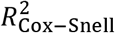 (15% of the maximum 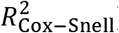) was used as recommended [20]. Based on the above parameters, the minimum sample size required for developing a new model is 149,750 for men and 299,750 for women. Hence, the derivation dataset needs a minimum sample of about 450,000 men and women.

With over 9 million patients in the open cohort and an estimated 7,000 incident liver cancer cases during 2008-2018 in the QResearch database, there is sufficient data for the development and validation datasets. We will use all the eligible patients in the database to maximise the power.

#### Exploration of non-linear relationships

Before imputation, a complete-case analysis will be fitted using a model containing only the continuous variables (e.g. age, BMI) within the development dataset to derive the fractional polynomial terms (up to two polynomial terms) [22, 23] for the non-linear relationships. Then a MICE model will be fitted in the development dataset to impute missing values and will include all predictors along with age interaction terms, the Nelson-Aalen estimator of the baseline cumulative hazard, and the outcome indicator (namely, incident liver cancer). Separate models will be fitted for men and women.

#### Model development

We will use similar established analytical strategies to develop and evaluate the risk prediction equations in this study that were used in the other QResearch studies [8, 24-28]. The internal-external validation approach [29, 30] will be used to quantify the heterogeneity of predictor effects in different geographical regions in England, rather than splitting data by general practice into development and validation datasets. Eight geographical regions will be used to develop the model and the remaining two regions to validate the model. This process will be performed five times, leaving out two different regions at each time. The indicators of model performance (subsection below) from the internal-external validation approach will be pooled using random-effects meta-analysis [29, 31]. The final model will be used to derive risk estimates for each year of follow-up, with a specific focus on 1-year (short-term), 5-year (medium-term) and 10-year (long-term) risk estimates. Separate models will be developed and evaluated for men and women, as the coefficients for the risk factors may be different between sexes.

Cox proportional hazards model will be used as the main method to develop risk prediction models, using robust variance estimates to allow for clustering of patients within general practices, also accounting for censoring (due to death, loss of follow-up, or the end of the observation period). The proportional hazards assumptions for Cox regression will be checked. The time origin is the date of entry into the study cohort. The risk period of interest is from the time origin up to the date of incident diagnosis of liver cancer. Patients who died from other causes will be considered as censored. The main analyses will be multivariable models including various predictors and interaction terms. The regression coefficients for each variable in the final model will be used as weights. From which, we will use a formula to derive the absolute risk estimates by combining them with the baseline survivor function evaluated for each year of follow-up, with a maximum of 10 years [32]. We will compare our developed model with the other validated risk prediction models in this field [33].

#### Model validation – evaluate the model performance

An imputation model (MICE) will be fitted for missing values in the validation datasets with five imputations (same as in the deviation dataset) using the methods described in the earlier subsection. We will apply the risk equations for males and females derived from the previous step to the validation data and calculate measures of model performance.

As in previous studies [34], we will calculate the R^2^ [35], the D statistic [36], the Brier score [37], and Harrell’s C statistics [38] at 1, 5, and 10 years and combine these across the imputed datasets using Rubin’s rules. R^2^ is the explained variation, where a higher value indicates a greater proportion of variation in survival time is explained by the model [35]. The D statistic is a measure of discrimination, which quantifies the separation in survival between patients with different levels of predicted risk, where higher values indicate better discrimination [36]. The Brier score is an aggregate measure of disagreement (the average squared error difference) between the observed and the predicted outcomes [37]. The Harrell’s C statistic [38] is a measure of discrimination (separation) that quantifies the extent to which those with earlier events have higher risk scores. Higher values of Harrell’s C indicate better performance of the model for predicting the relevant outcome. A value of 1 indicates that the model has perfect discrimination. A value of 0.5 indicates that the model discrimination is no better than chance. The 95% confidence intervals for the performance statistics will be calculated to allow comparisons with alternative models for the same outcome and across different subgroups [39].

We will assess the calibration of the risk scores by comparing the mean predicted risks at 10 years with the observed risks by categories of the predicted risk (e.g. by decile or twentieth), which will be presented in a calibration plot. The observed risks for men and women can be obtained by using the Kaplan-Meier estimates. We will also evaluate these performance measures in six pre-specified age groups (25-39, 40-49, 50-59, 60-69, 70-79, 80+). Decision curve analysis [40] will be used to evaluate the net benefit of the prediction model (clinical usefulness). We will follow the recommendations from the TRIPOD guideline [17] to report the multivariable prognostic model.

#### Risk stratification

Risk stratification allows patients with a high predicted risk to be identified electronically from primary care records for tailored advice, active monitoring of the disease progression, and screening. Since there is no widely accepted threshold for classifying a high risk of liver cancer, we will examine the distribution of the predicted risks and calculate a series of centile values in the model. For each centile threshold, we will calculate the sensitivity and specificity of the risk scores. The currently accepted threshold for classifying high risk in other QCancer models is 3% [41].

#### Dissemination and implementation plan of the prediction model

The developed algorithm will be published in peer-reviewed journals and presented at academic conferences. A web-based program can implement the new risk algorithm similar to the QCancer tool (https://www.qcancer.org/), subject to funding and the Medicines and Healthcare Products Regulatory Agency (MHRA) medical device compliance. It will also be possible to implement the risk algorithm in the EHR systems, using existing data to calculate individual risks for the primary care population. These implementation intentions will be subject to the terms and conditions of QResearch, the University of Oxford, the Cancer Research UK grant, and the agreement of all parties. It will be covered by another implementation protocol, which is out of the scope of this research protocol.

### Summary: relevant guidelines used in this project

- NICE NG12 (Suspected cancer: recognition and referral) [41]
- The European Association for the Study of the Liver (EASL) clinical practice guidelines (management of HCC) [42]
- The STROBE statement (reporting guideline for observational studies) [19]
- The TRIPOD statement (reporting guideline for multivariable prediction model) [17, 43]

## Discussion

### The strengths and limitations of this project

To our best knowledge, this study will be the largest of its kind for early detection and diagnosis of hepatocellular liver cancer from the English primary care population. It has significant potential to address several important research gaps and gain a deeper understanding of the presentation, characteristics, and outcomes of liver cancer. The key strengths of this population-based study include prospective recording of predictors and outcomes, good ascertainment of outcomes through linkage of multiple national databases, and a large sample size from an established validated database (QResearch) that has been used to develop many risk prediction tools, such as QRisk3 [27], QCancer (10-year risk) [8] and other algorithms. UK primary care records have high levels of accuracy and completeness of clinical diagnoses and prescribed medications. This study has good face validity, as it is conducted in the same setting where most patients are clinically assessed, managed, and followed up in England. Thanks to the expansion and upgrade of the QResearch database in recent years, we now have information on cancer staging, grade, and histology. The rich data source makes in-depth exploration possible. This study also minimises the most common biases in epidemiological studies, such as selection bias, recall bias, and respondent bias. We will also use relevant clinical and statistical guidelines to follow the best research practice and to guide the analysis and report the findings for transparent and reproducible research. All of these strengths make the study findings more robust and more likely to generalise to the wider UK population.

The limitations of this study may include potential information bias and missing data. Based on our experiences of using primary care data, some lifestyle factors such as BMI, smoking and alcohol status may not always track the true values in real-time, and often lack consistency. In addition, the recording of family history of cancer in primary care records may be sparse. Although the completeness of cancer staging is improving year by year, we are concerned that incomplete cancer staging limits further exploration of the factors influencing early/late diagnosis of liver cancer. However, we may overcome this limitation by imputing cancer stage, as we have rich clinical data, treatments, and survival outcomes linked to the QResearch database, which can be used in the imputation model.

Due to the available resources and the funding, we will use the internal-external approach to validate the developed model using data from the same database. QResearch uses data from the EMIS, which is the computer system used by 55% of the GP surgeries in the UK. Our study population is based in England and representative of the whole English primary care population. Some previous studies [44-46] independently examined other risk equations developed by the QResearch team and concluded that using external data gave a similar performance as the internal validation approach using the QResearch database, which is reassuring. It may be possible to externally validate the developed model by our collaborators in Scotland or Denmark using datasets outside of England in future studies.

### Clinical implications and research impact

This study will generate new knowledge regarding the risk factors, early signs and clinical features (symptoms and comorbidities) of liver cancer from the English primary care population. Such findings will allow us to gain a deeper understanding of the current situation of liver cancer in England, and have implications on health policy to early detect, diagnose, and manage liver cancer at the population level. Individuals at the highest risk of developing liver cancer are most likely to benefit from active surveillance (e.g. using liver ultrasonography and serum alpha-fetoprotein test). Applying personalised risk prediction models to select individuals at high risk from the population and referring them to specialists for further investigation could be a cost-effective approach to improve early diagnosis of liver cancer and patient outcomes, without unduly burdening the overstretched NHS, and avoiding surveillance harms for patients at low risk. The implementation of the prediction model will allow the public to calculate individual risks of developing liver cancer for free, like the other QCancer tools, which is a way to engage the public to be more aware of their health status and initiate help-seeking when necessary. In summary, this study has great potential of making contributions to the national early detection and diagnosis cancer strategic plan, with associated public and societal benefits (e.g. reduce premature death due to liver cancer, reduce care costs for the NHS, and reduce disease burden to the society).

## Supporting information

DeLIVER consortium authors

## Data Availability

Considering the sensitive nature of anonymised patient-level data and to meet the requirement of the data provider, the data will be only accessible to the named researchers of the team.

## Declarations

### Authors’ contributions

EB and JH-C secured the funding. EB is the chief investigator of the DeLIVER project, and JH-C is the package lead and the guarantor of this study. All authors contributed to the study conceptualisation. WL specified the data, led on the ethical approval, and is the lead statistician for the DeLIVER-QResearch project. WL designed the statistical analysis plan, with methodological input from JH-C, C. Coupland, PJ and HI, clinical input from PJ, JH-C, EB and PM, and contextual input from JH-C, PJ, EB, PM, HI and C. Campbell. WL drafted the whole research protocol. All authors read and commented on the earlier drafts, contributed to the revision of the manuscript, and approved the final version of the manuscript for submission.

## Acknowledgements

We are grateful that two named patient representatives (both diagnosed with HCC) reviewed our lay summary for ethical approval and provided very helpful feedback. They also commented on the list of potential symptoms and comorbidities of liver cancer that we provided, and shared their disease trajectories and personal lived experiences of HCC with us. We also thank the former project manager, Dr Katja Pfafferott, for coordinating the patient and public involvement (PPI) process and the project setup.

This project involves anonymised data from patient-level information collected by the NHS, as part of the care and support of patients. We acknowledge the contribution of the patients and general practices contributing to the EMIS (Egton Medical Information Systems) clinical computer system and the QResearch database, and the Universities of Nottingham and Oxford for the expertise in establishing, developing, and supporting the QResearch database. The cancer registration data used in this study are supplied by Public Health England. The civil (mortality) registration data are provided by the Office for National Statistics. The Hospital Episode Statistics data used in this study are re-used with permission from NHS Digital, who retain the copyright of the data. None of the acknowledged organisations or funding bodies has been involved in any research process, including study design, data specification, statistical analysis, interpretation of results, preparing manuscripts, or decision to publish.

## Study and authors’ information

DeLIVER website: www.deliver.cancer.ox.ac.uk

## Twitter

@DeLIVER_HCC (project), @WLiao_Ox (Weiqi Liao), @pippa_matt (Philippa Matthews),

@EleanorBarnesOx (Eleanor Barnes), @JuliaHCox (Julia Hippisley-Cox), @QResearchOxford (QResearch)

## Co-investigators and members in the DeLIVER consortium

Please refer to the appended file.

