## Supplementary material for "Development and validation of personalised risk prediction models for early detection and diagnosis of primary liver cancer among the English primary care population using the QResearch^®^ database: research protocol and statistical analysis plan": DeLIVER consortium authors

| | Full name | Email Address | Primary affiliation | Secondary affiliation | ORCID ID | Funding acknowledgements | Other acknowledgements |
|---|---|---|---|---|---|---|---|
| 1 | Eleanor Barnes | | Niuffield Department of Medicine | OUH NHS trust | 0000-0002-0860-0831 | This work was supported by Cancer Research UK (C30358/A29725). EB is supported by the Oxford NIHR Biomedical Research Centre and is an NIHR Senior Investigator. | The views expressed in this article are those of the author and not necessarily those of the NHS, the NIHR, or the Department of health. |
| 2 | Emma Culver | | Translational Gastroenterology Unit, JRH Oxford | NDM, University of Oxford. | 0000-0001-9644-8392 | BRC Oxford | |
| 3 | Roman Fischer | | Target Discovery Institute/NDM | CAMS Oxford Institute (COI) | 0000-0002-9715-5951 | COI | |
| 4 | Julia Hippisley-Cox | | Nuffield Department of Primary Care Health Sciences, University of Oxford | | http://orcid.org/0000-0002-2479-7283 | Funding from CRUK Oxford centre, John Fell fund, ISSF wellcome for the QResearch | |
| 5 | Hamish Innes | | Glasgow Caledonian University; School of Health and Life Sciences. | University of Nottingham; Division of Epidemiology and Public Health TERTIARY AFFILIATION: Public Health Scotland; Glasgow. | | Medical Research Foundation (Grant ID: C0825) | |
| 6 | William L Irving | | NIHR Nottingham Biomedical Research Centre, Nottingham University Hospitals NHS Trust and the University of Nottingham | | 0000-0002-7268-3168 | | |
| 7 | Peter Jepsen | | Department of Hepatology and Gastroenterology, Aarhus University Hospital, Aarhus, Denmark. | | 0000-0002-6641-1430 | Peter Jepsen's work is funded by a grant from the Novo Nordisk Foundation. Grant reference number: NNF19OC0054612. | |
| 8 | Matt Kelly | | Perspectum | | 0000-0002-5834-635X | Perspectum employee. | |
| 9 | Paul Klenerman | | Nuffield Department of Medicine | Translational Gastroenterology Unit | 0000-0003-4307-9161 | supported by Oxford NIHR BRC | |
| 10 | Weiqi Liao | | Nuffield Department of Primary Care Health Sciences, University of Oxford | | https://orcid.org/0000-0002-8605-3749 | | |
| 11 | Derek Mann | | Newcastle University | Biosciences Institute | https://orcid.org/0000-0003-0950-243X?lang=en | CRUK and MRC programme grants and CRUK HUNTER consortium | |
| 12 | Dr Aileen Marshall | | Royal Free Hospital, London | Institute of Liver and Digestive Health, UCL | 0000-0003-3283-6315 | None at present. NHS staff. | |
| 13 | Philippa C Matthews | philippa.matthews@ | The Francis Crick Institute, 1 Midland Rd, London | University College London | 0000-0002-4036-4269 | Wellcome Grant Ref 110110/Z/15/C, core funding from the Francis Crick Institute and University College London NIHR BRC | University College London NIHR BRC |
| 14 | Michael Pavlides | | Radcliffe Department of Medicine | Translational Gastroenterology Unit | https://orcid.org/0000-0001-9882-8874 | supported by Oxford NIHR BRC | |
| 15 | Rory J R Peters | | Nuffield Department of Medicine | Translational Gastroenterology Unit | 0000-0003-4347-9739 | CRUK Clinical Research Training Fellowship | |
| 16 | Elisabeth Pickles | | Institute of Biomedical Engineering, University of Oxford | Perspectum | 0000-0003-4974-4943 | Royal Commission for the Exhibition of 1851 Industrial Fellowship | |
| 17 | James Robineau | | University of Oxford | | | | |

| | Full name | Email Address | Primary affiliation | Secondary affiliation | ORCID ID | Funding acknowledgements | Other acknowledgements |
|---|---|---|---|---|---|---|---|
| 18 | Benjamin Schuster-Böckler | | Nuffield Department of Medicine | Ludwig Institute for Cancer Research | 0000-0002-8892-5133 | supported by Ludwig Cancer Research | |
| 19 | Chunxiao Song | | Ludwig Institute for Cancer Research, Nuffield Department of Medicine, University of Oxford, Oxford, UK. | Target Discovery Institute, Nuffield Department of Medicine, University of Oxford, Oxford, UK. | 0000-0002-7781-6521 | Ludwig Institute for Cancer Research, Cancer Research UK (C63763/A26394 and C63763/A27122), National Institute for Health Research (NIHR) Oxford Biomedical Research Centre (BRC), Emerson Collective | |
| 20 | Jeremy Tomlinson | | Radcliffe Department of Medicine | OCDEM | 0000-0002-3170-8533 | supported by Oxford NIHR BRC | |
| 21 | Christopher Welberry | | Oncimmune ltd, Nottingham, United Kingdom | N/A | 0000-0001-7432-3361 | N/A | N/A |

Note: The co-investigators and members in the DeLIVER consortium are **in alphabetical order by surname** in this table.
